# Comparing COVID-19 risk factors in Brazil using machine learning: the importance of socioeconomic, demographic and structural factors

**DOI:** 10.1101/2021.03.11.21253380

**Authors:** Pedro Baqui, Valerio Marra, Ahmed M. Alaa, Ioana Bica, Ari Ercole, Mihaela van der Schaar

## Abstract

**Background:** The COVID-19 pandemic continues to have a devastating impact on Brazil. Brazil’s social, health and economic crises are aggravated by strong societal inequities and persisting political disarray. This complex scenario motivates careful study of the clinical, socioeconomic, demographic and structural factors contributing to increased risk of mortality from SARS-CoV-2 in Brazil specifically.

**Methods:** We consider the Brazilian SIVEP-Gripe catalog, a very rich respiratory infection dataset which allows us to estimate the importance of several non-laboratorial and socio-geographic factors on COVID-19 mortality. We analyze the catalog using machine learning algorithms to account for likely complex interdependence between metrics.

**Findings:** The XGBoost algorithm achieved excellent performance, producing an AUC-ROC of 0.813 (95%CI 0.810–0.817), and outperforming logistic regression. Using our model we found that, in Brazil, socioeconomic, geographical and structural factors are more important than individual comorbidities. Particularly important factors were: The state of residence and its development index; the distance to the hospital (especially for rural and less developed areas); the level of education; hospital funding model and strain. Ethnicity is also confirmed to be more important than comorbidities but less than the aforementioned factors.

**Interpretation:** Socioeconomic and structural factors are as important as biological factors in determining the outcome of COVID-19. This has important consequences for policy making, especially on vaccination/non-pharmacological preventative measures, hospital management and healthcare network organization.

**Funding:** None.

## INTRODUCTION

The COVID-19 pandemic is having a particularly devastating impact on Brazil with, at the time of writing, over 270,000 registered cumulative deaths, second only to the USA.^1^ Brazil’s social, health and economic crises are aggravated by strong societal inequities^2^ and political disarray.^3^ COVID-19 outcomes are likely to be the result of the interplay between patient and environmental factors. Age is now well established as the dominant determinant of mortality.^4^ We have previously demonstrated the important effect of ethnicity and socioeconomic status in determining outcome in Brazil.^2^ A number of institutional and organizational effects may also be important. It has been shown that treatment site seems to have a substantial association with mortality comparable to the effect of co-morbidity, at least for intensive care outcomes.^5^ This suggests that institutional and organizational factors may be important. This is reasonable as it is likely that different hospitals may vary in their ability to respond to a surge in cases either because they are locally overwhelmed, experience an early influx of patients before surge capacity can be put into place or because they are inherently less able to expand capacity. A limited number of studies have attempted to look at this. A recent study in the United States did find evidence to support an association between hospital strain and increased mortality^6^ for critical care patients, but not ward patients, and that this relationship changed over time. A similar negative impact of intensive care capacity was seen in Belgium.^7^ A full understanding of the interplay between patient and healthcare system factors is crucial for rational, dynamic allocation of hospital resources as well as the targeting of both pharmacological and non-pharmacological interventions. Healthcare systems vary substantially around the world, making local evaluation important. To our knowledge, this has not previously been undertaken in Brazil. Healthcare organizational factors are likely to be, to some extent, co-linear with other socioeconomic predictors and their effects may be non-linear: The extent to which organizational effects are real or the result of a failure to completely adjust for other factors in a linear model is not known. This observation motivates the use of explainable machine learning models able to deal with complex interactions and non-linear relationships.

### Research in context

#### Evidence before this study

Brazil has been particularly badly affected by COVID-19, experiencing one of the highest death tolls globally. The country has a complex socioeconomic and ethnic composition and we have previously shown that these considerations are an important determinant of outcome. To date, however, there has not been a study of other structural factors that may contribute to mortality in Brazil. We use a machine learning approach to automatically handle likely complex interdependencies between a large number of features of interest.

#### Added value of this study

We confirm the effects of ethnicity, socioeconomic status and age on outcome. We show that, accounting for age (and therefore accounting for the dominant clinical determinant), mortality probability distributions are differently skewed favoring survival in privately funded hospitals in richer areas, particularly for younger patients. Additionally we show that structural factors such as distance to hospital and an index of hospital strain are as important as comorbidities.

#### Implications of all the available evidence

Our research shows that mortality is highest in poorer, rural areas where hospital are distant or are overwhelmed. This may have important policy implications in rational configuration of both surge capacity as well as other pharmacological or non-pharmacological measures where this is not possible.

In this study, we use the Brazilian SIVEP-Gripe respiratory infection surveillance dataset^8^ to study demographic, patient, socioeconomic and organizational structure influences on COVID-19 outcome. As depicted in Figure 1, we model the linear and nonlinear correlations among the covariates using the successful XGBoost machine learning technique. We name ‘XCOVID-BR’ the XGBoost model that achieves the highest performance. The goal of this work is to provide the scientific community and, in particular, the Brazilian authorities with a ranking of the most important social, health and economic risk factors.

**Figure 1.**
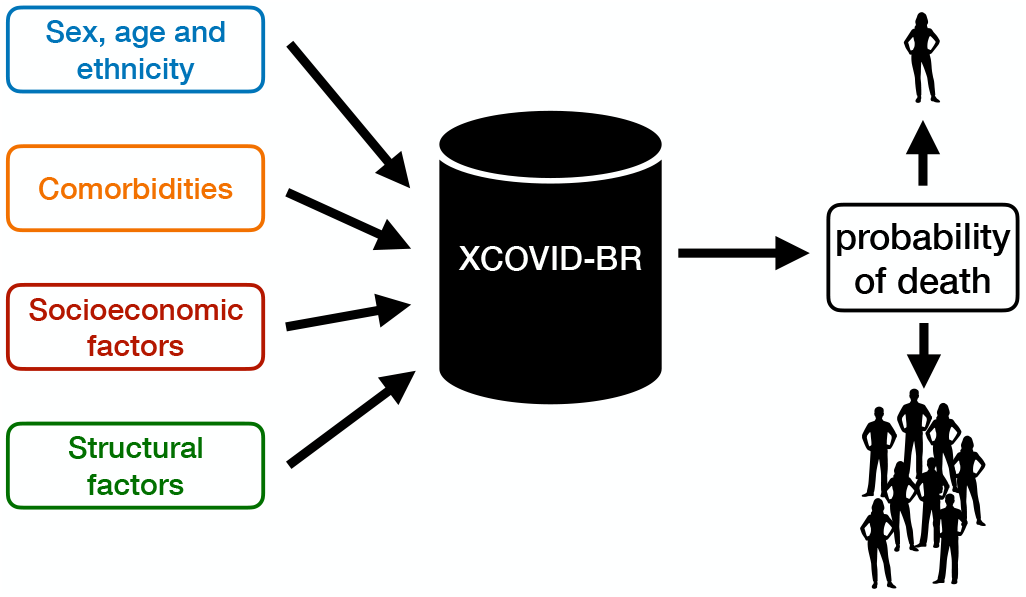
The XCOVID-BR machine learning model. XCOVID-BR takes as input a range of medical, socioeconomic and structural factors and returns as output the probability of death by COVID-19. XCOVID-BR can be applied to individuals, groups or whole sections of the Brazilian population.

## METHODS

We analyze COVID-19 hospital mortality using the public SIVEP-Gripe dataset (Sistema de Informação de Vigilância Epidemiológica da Gripe), a prospectively collected respiratory infection registry which is maintained by the Brazilian Ministry of Health for the purposes of recording cases of severe acute respiratory syndrome (SARS) across both public and private hospitals.^8^ We analyze data collected from February 25 to September 21, 2020. Out of the 279,987 hospitalized patients that had a positive RT-PCR test for SARS-CoV-2, 242,679 cases have known outcome and age *≤* 110. We consider only patients who were admitted to hospital in order to be less sensitive to the regional variability of testing. Finally, as we are interested also in socioeconomic factors, we restrict our analysis to the 231,112 patients whose files contain geographic information and type of health-care (public or private). See Figure 2.

**Figure 2.**
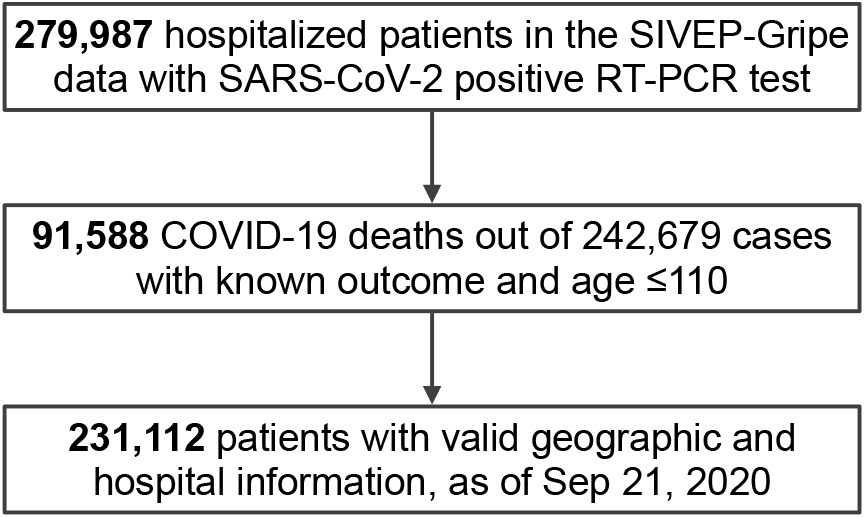
Flowchart of SIVEP-Gripe data used in this study. SARS-CoV-2 stands for severe acute respiratory syndrome coronavirus 2. SIVEP-Gripe stands for *Sistema de Informação de Vigilância Epidemiológica da Gripe*.

We initially consider 30 patient features including clinical (age, sex, ethnicity, comorbidities and symptoms), socio-geographic (education, state, municipal human development index MHDI, city type) and structural hospital-level (distance from patient to hospital, time-dependent strain and funding) factors. In order to capture the time-varying pressure on individual hospitals, we defined ‘hospital strain’ as the number of hospitalized patients during the admission week divided by a metric of hospital capacity. As capacity numbers data were not available for all the hospitals considered, we used as a proxy the total number of hospitalizations according to the 2019 SIVEP-Gripe dataset. The 231,112 patients that we consider come from 1,801 different cities and from 3,991 different hospitals. This abundance and richness allows us to disentangle the importance of a factor from the one of its covariates, fully considering all the correlations.

The prediction task was formulated as a binary classification problem for hospital mortality, with 0 representing death and 1 representing recovery. The analysis was performed using XGB but Logistic Regression, K-Nearest Neighbors, Neural Network, Random Forest and Support Vector Machine algorithms were also evaluated and are included in the Supplementary Materials for completeness. Our models are implemented in Python through the scikit-learn and XGB packages.^9,10^

For the training and test sets, we used 80% (184,889 patients) and 20% (46,223 patients) random split with *k*=10-fold cross validation. As metrics we consider the area under the receiver-operating characteristic curve (AUC-ROC) and the average precision (AP), which is the area under the precision-recall curve relative to a given classification (recovery or death). Feature importance is analyzed using the permutation method (see the Supplementary Materials for more details and robustness tests).

The SIVEP-Gripe catalog has missing values. In the case of comorbidities or symptoms we imputed missing values as the clinical feature being absent for the individual.^2^ For the remaining variables we did not perform pre-processing for the XGB algorithm as the latter already imputes missing data. A table with the percentages of patients with missing values is available in the Supplementary Materials.

The study was conducted and reported in line with the Transparent Reporting of a multivariable prediction model for Individual Prognosis Or Diagnosis (TRIPOD).^11^ The TRIPOD Statement is available in the Supplementary Materials.

## RESULTS

Our ‘XCOVID-BR’ XGBoost model achieved the highest performance of the models considered (Supplementary Materials) and is considered in the subsequent analysis. The model is publicly available at github.com/PedroBaqui/XCOVID-BR. We found that the XGBoost algorithm achieves excellent performance of AUC-ROC=0.813 (95%CI 0.810–0.817) compared to the logistic regression’s AUC-ROC=0.766 (95%CI 0.761– 0.770, full comparison table in the Supplementary Materials). Model calibration is shown in Figure 3.

**Figure 3.**
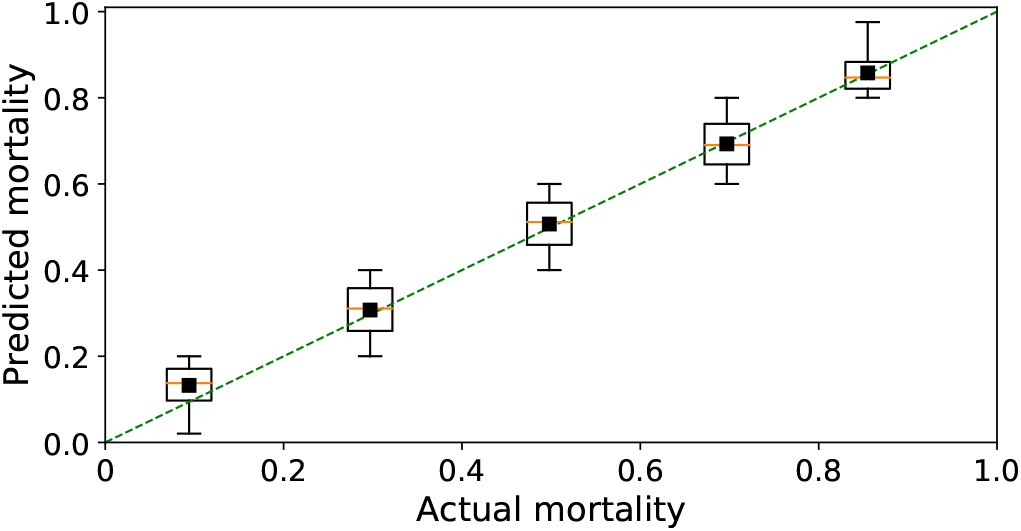
Model calibration. The calibration analysis shows that mortality predicted by the XCOVID-BR model performs uniformly across bins of mortality.

Figure 4 shows the importance of the features considered in this analysis, excluding symptoms as they are not related to the patients’ pre-infection conditions from the XCOVID-BR model.

**Figure 4.**
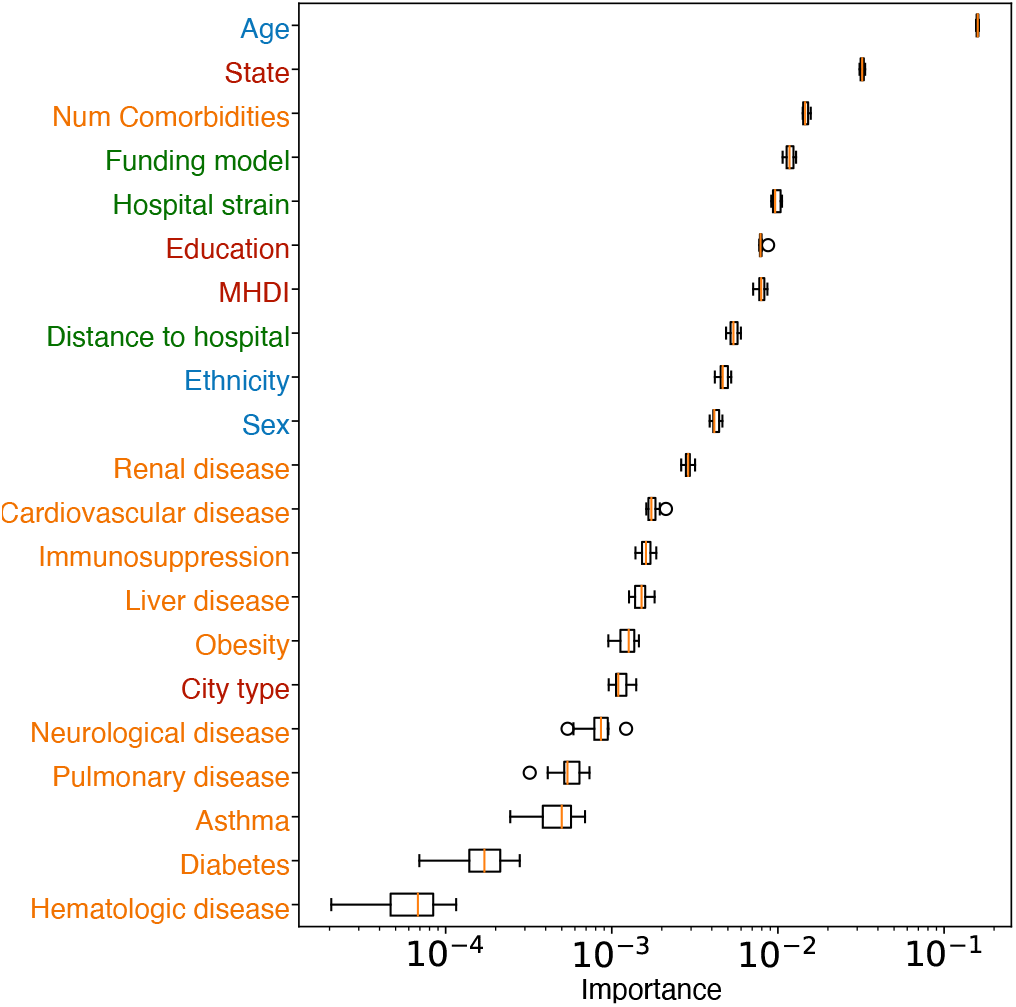
Relative feature importance (median and IQR) for mortality risk to COVID-19. The coloring marks the categories listed in Figure 1. Feature importance was estimated via the permutation method and a logarithmic scale is employed for clarity.

In Figure 5 we again use the permutation method, but we split the test set between younger (<60 years, AUC-ROC=0.770, 95%CI 0.763–0.777) and older (*≥* 60 years, AUC-ROC=0.717, 95%CI 0.711–0.725) patients, 24,277 and 21,946 patients respectively (in Brazil patients over 60 are considered elderly^2^). We find that for younger patients the state and the number of comorbidities play a more important role than their own age (Figure 5). On the other hand, for elderly patients state has a greater importance and variations in age within this group are disproportionately important.

**Figure 5.**
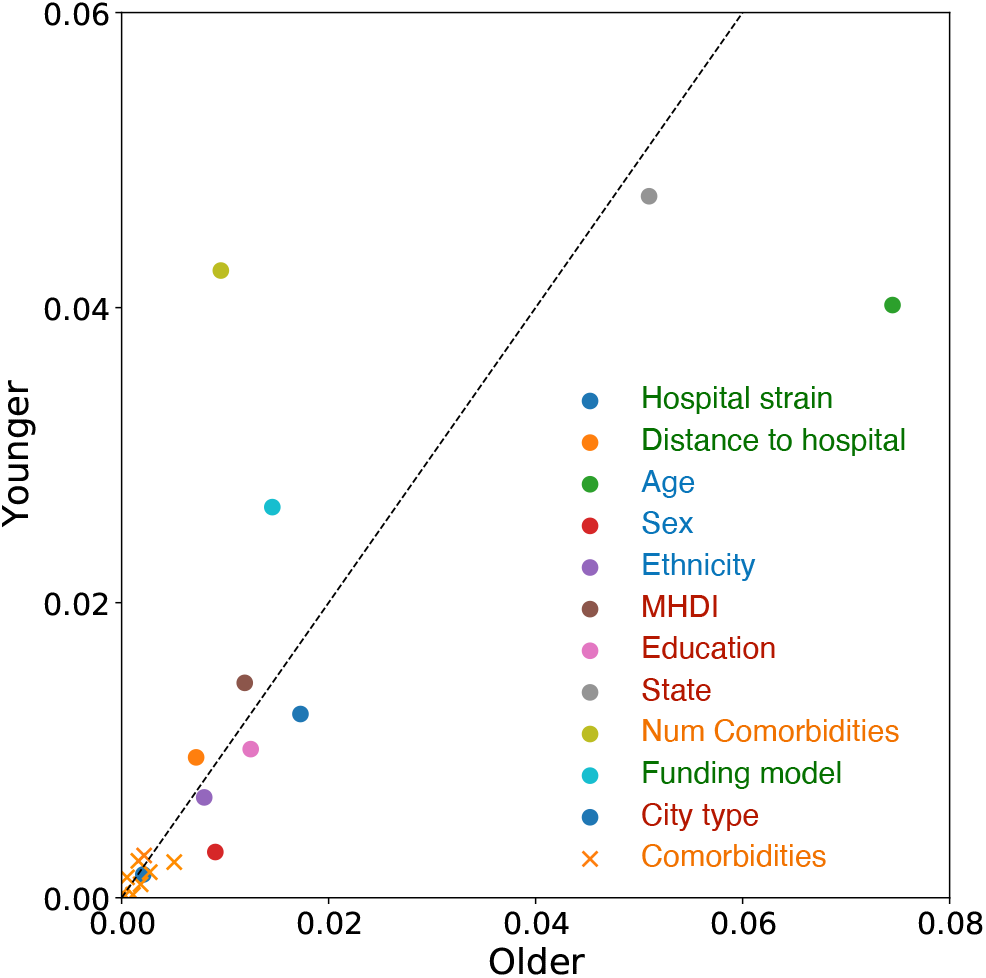
Feature importance for older and younger patients. Each point represents a feature in the SIVEP-Gripe dataset, ordered by the importance for COVID-19 mortality prediction for older (*≥* 60 years) and younger (<60 years) hospitalized patients. Variables deviating from the dotted identity line suggests a different relative importance for the groups. The coloring marks the categories listed in Figure 1.

As seen from the analysis of Figure 4, hospital funding model (private or public) is an important feature. Figure 6 shows the mortality rate for patients admitted to publicly and privately hospitals, stratified according to age (the dominant factor associated with outcome).

**Figure 6.**
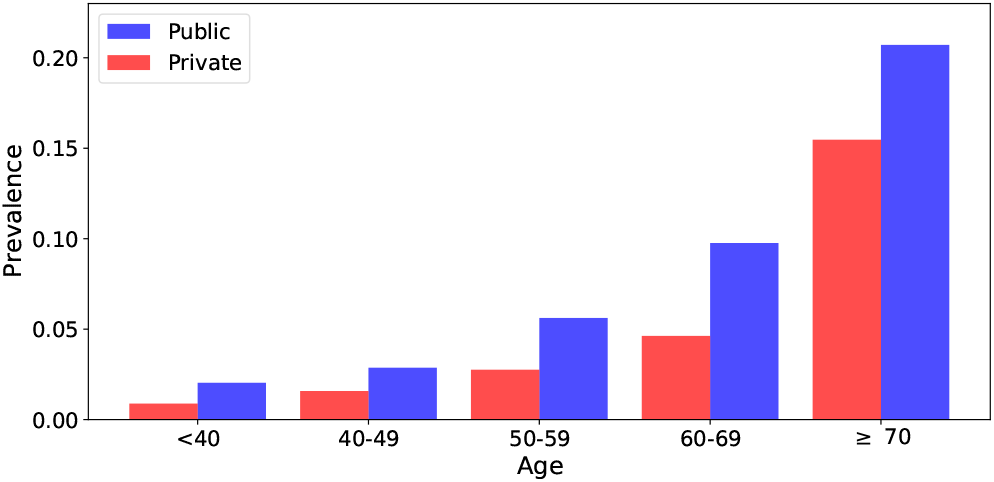
Mortality rate for public and privately funded hospitals, stratified according to age. The bars are normalized by dividing the fatalities by the total number of cases for each type of hospital funding model.

Table I shows the demographic and socio-geographic characteristics and coexisting conditions among survivors and non-survivors. We also show the AUC-ROC relative to the XGB algorithm for patients belonging to each category.

**Table I.** Demographic and socio-geographic characteristics and coexisting conditions among survivors and non-survivors of COVID-19. Data are *n* (%) or mean (SD). The last column gives the AUC-ROC relative to the XGB algorithm for patients belonging to each category.

|  | Survivors | Non-survivors | AUC (95%CI) |
| --- | --- | --- | --- |
| Brazil | 143889 (62.3%) | 87223 (37.7%) | 0.813 [0.810, 0.817] |
| Age (years) | 54.3 ± 17.7 | 68.6 ± 15.5 | – |
| Sex |  |  |  |
| Women | 63919 (63.5%) | 36700 (36.5%) | 0.811 [0.805, 0.816] |
| Men | 79941 (61.3%) | 50511 (38.7%) | 0.815 [0.810, 0.820] |
| Funding model |  |  |  |
| Private | 35830 (74.7%) | 12148 (25.3%) | 0.843 [0.832, 0.851] |
| Public | 108059 (59.0%) | 75075 (41.0%) | 0.799 [0.795, 0.802] |
| MHDI |  |  |  |
| Very High | 63888 (70.0%) | 27430 (30.0%) | 0.816 [0.810, 0.823] |
| High | 72657 (58.5%) | 51552 (41.5%) | 0.802 [0.797, 0.808] |
| Medium-Low | 7344 (47.1%) | 8241 (52.9%) | 0.791 [0.776, 0.809] |
| Ethnic group |  |  |  |
| White | 55040 (64.1%) | 30874 (35.9%) | 0.813 [0.806, 0.821] |
| Black | 6585 (57.5%) | 4871 (42.5%) | 0.803 [0.780, 0.816] |
| East Asian | 1521 (60.2%) | 1005 (39.8%) | 0.789 [0.742, 0.825] |
| Brown | 38700 (57.0%) | 29205 (43.0%) | 0.804 [0.797, 0.812] |
| Indigenous | 280 (56.6%) | 215 (43.4%) | 0.891 [0.821, 0.948] |
| Macro-regions |  |  |  |
| North | 5974 (49.1%) | 6192 (50.9%) | 0.781 [0.765, 0.796] |
| Northeast | 21830 (51.7%) | 20421 (48.3%) | 0.800 [0.791, 0.807] |
| Central-West | 11877 (63.9%) | 6724 (36.4%) | 0.810 [0.796, 0.824] |
| Southeast | 84551 (65.4%) | 44796 (34.6%) | 0.811 [0.805, 0.816] |
| South | 19657 (68.4%) | 9090 (31.6%) | 0.809 [0.800, 0.822] |
| City type |  |  |  |
| Urban | 124635 (62.6%) | 74523 (37.4%) | 0.813 [0.808, 0.818] |
| Peri-Urban | 382 (54.7%) | 316 (45.3%) | 0.795 [0.711, 0.859] |
| Rural | 3288 (54.0%) | 2797 (46.0%) | 0.798 [0.769, 0.823] |
| Education level |  |  |  |
| Illiterate | 2048 (39.3%) | 3169 (60.7%) | 0.762 [0.733, 0.790] |
| Elem. school I | 10912 (51.0%) | 10499 (49.0%) | 0.773 [0.759, 0.785] |
| Elem. school II | 9026 (59.1%) | 6242 (40.9%) | 0.808 [0.793, 0.824] |
| High school | 18244 (71.0%) | 7459 (29.0%) | 0.828 [0.817, 0.840] |
| Higher education | 9603 (77.6%) | 2778 (22.4%) | 0.874 [0.860, 0.889] |
| Comorbidities |  |  |  |
| Cardiovascular d. | 43068 (53.1%) | 37983 (46.9%) | 0.767 [0.760, 0.774] |
| Asthma | 4453 (68.8%) | 2021 (31.2%) | 0.810 [0.781, 0.837] |
| Diabetes | 31719 (52.2%) | 29046 (47.8%) | 0.758 [0.750, 0.766] |
| Pulmonary dis. | 4007 (42.1%) | 5500 (57.9%) | 0.746 [0.723, 0.772] |
| Obesity | 7161 (59.0%) | 4970 (41.0%) | 0.771 [0.754, 0.793] |
| Immunosuppr. | 3407 (47.3%) | 3795 (52.7%) | 0.741 [0.715, 0.766] |
| Renal dis. | 4112 (38.3%) | 6632 (61.7%) | 0.719 [0.699, 0.738] |
| Liver dis. | 925 (40.2%) | 1377 (59.8%) | 0.731 [0.695, 0.773] |
| Neurological dis. | 3905 (39.2%) | 6068 (60.8%) | 0.727 [0.701, 0.753] |
| Hematologic dis. | 1054 (50.5%) | 1035 (49.5%) | 0.804 [0.767, 0.851] |
| Symptoms |  |  |  |
| Fever | 99656 (65.1%) | 53497 (34.9%) | 0.813 [0.808, 0.818] |
| Vomiting | 12269 (67.0%) | 6049 (33.0%) | 0.810 [0.793, 0.826] |
| Cough | 110183 (65.0%) | 59246 (35.0%) | 0.808 [0.804, 0.813] |
| Sore throat | 28238 (68.5%) | 12979 (31.5%) | 0.821 [0.812, 0.831] |
| Respiratory disc. | 74879 (56.8%) | 56912 (43.2%) | 0.803 [0.798, 0.808] |
| Shortness breath | 96954 (58.8%) | 68033 (41.2%) | 0.802 [0.797, 0.806] |
| Diarrhea | 21837 (69.1%) | 9746 (30.9%) | 0.811 [0.799, 0.820] |
| SpO<sub>2</sub> < 95% | 75083 (55.2%) | 61021 (44.8%) | 0.787 [0.780, 0.795] |

Finally, we adopt the XCOVID-BR model in order to estimate the mortality risk of specific sections of the Brazilian population. Given a patient’s non-laboratorial data, the XCOVID-BR model returns a probability ranging from 0 (death) to 1 (recovery). One can then estimate the overall risk of a group by studying the distribution of the XCOVID-BR outcomes. Figure 7 shows the XCOVID-BR model applied to age and hospital sub-groups taken from the states of Pernambuco (Northeast) and Paraná (South).

**Figure 7.**
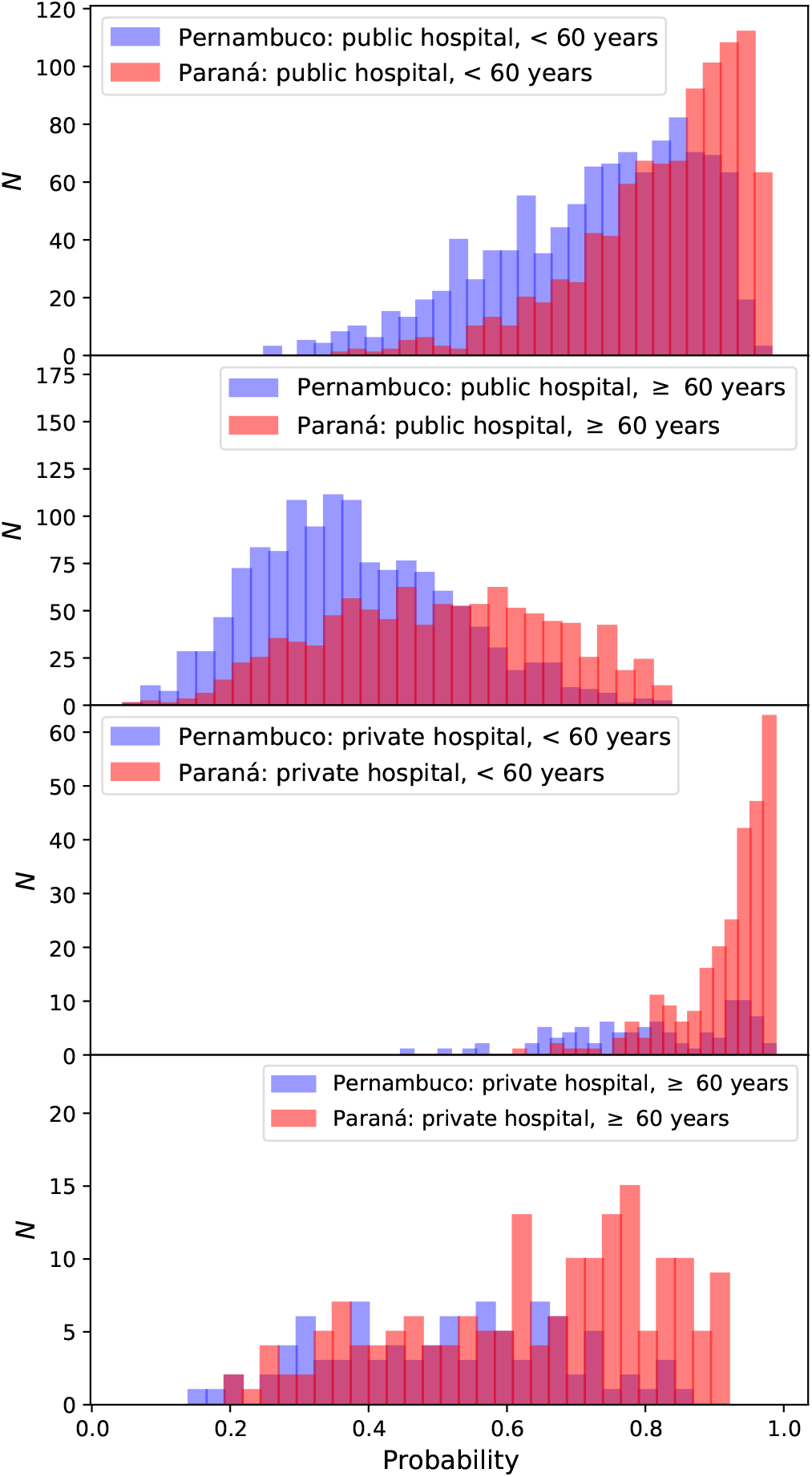
Representative probability of survival distributions as estimated by the XCOVID-BR model. We contrast typical publicly and privately funded hospitals from Pernambuco, an example of a region in the more socioeconomically challenged Northeast, with examples from the richer Paraná region in the South. Stratifying by age, the dominant *clinical* predictor of mortality, it is apparent that the probability distribution is skewed with lower mortality in the wealthier (Paraná) region and this is particularly apparent in younger patients and in privately-funded hospitals.

## DISCUSSION

We present, to our knowledge, the most extensive application of machine learning to COVID-19 hospital survival in Brazil. We considered the very rich SIVEP-Gripe dataset as of September 21, 2020. We confirm several worldwide findings but also report important sociodemo-graphic trends specific to Brazil.

We found that XGBoost outperforms other methods including logistic regression (Supplementary Materials). This improved performance demonstrates the non-linearity and co-linearity present in the data and justifies the choice of a machine learning model over conventional statistical techniques. The trained model is publicly available at github.com/PedroBaqui/XCOVID-BR.

Using XCOVID-BR we find that socioeconomic factors are more important than comorbidities (Figures 4 and 5), a scenario that seems to reflect the social inequalities present throughout Brazil. The number of comorbidities remains, however, the third most important feature, signaling that the interaction between comorbidities is a significant factor for the outcome of COVID-19 patients. We highlight the following factors: the state of residence and its development index, the distance to the hospital (very important for rural and less developed areas), the level of education, and hospital funding and strain. Ethnicity is also confirmed to be more important than comorbidities in agreement with an earlier investigation that adopted mixed-effects Cox regression survival analysis.^2^ Here, we also include socio-geographic features and model non-linear interactions via XGBoost and find that socio-geographic features are more important than ethnicity.

These findings also agree with the results from the descriptive analysis (Table I): non-survivors are older, more likely to have been admitted to public hospitals and live in less developed cities. Survivors are more likely to be white Brazilians, with high/higher education, living in urban areas. We also confirm the higher proportion of non-survivors in the North and Northeast macro-regions.^2^ Additionally, comorbidities, except for asthma, are more prevalent among non-survivors, especially renal and neurological diseases. The most common comorbidities were cardiovascular disease and diabetes.

Of course many of these variables are correlated: Patients with access to private healthcare tend to have a higher education and better living conditions (city development index). While the latter means that one shares a household with fewer people and the ready availability of basic services such as running water and sanitation, the former gives the possibility to work remotely. Poor literacy is likely to also impact negatively on healthcare access. These findings support the conclusion that socioeconomic, ethnic and geographical factors are crucial in order to correctly understand the pandemic in Brazil and plan adequate measures.^12^

We tested the predictive performance of the XCOVID-BR model for various sub-groups and found that the performance is generally similar to the global one, except for a few cases such as the North macro-region, illiterate Brazilians and some groups with comorbidities. The lower performance relative to these sub-groups indicates that it is more difficult to forecast the evolution of the disease within certain sections of Brazilian society, possibly because of higher heterogeneity within these sections. In other words, these groups are more susceptible to COVID-19, and it is also harder to study factors underlying their COVID-19 mortality risk. We hope this result will be useful in motivating federal authorities in adopting effective action in order to mitigate the impact of the pandemic for these groups.

Hospital funding model (private or public) was found to be a very important feature (Figure 4). We indeed clearly observe that public healthcare suffers from a higher mortality rate across all ages (Figure 6). This is not unexpected as private healthcare serves only 25% of the Brazilian population and total spending is similar to that of public healthcare, implying that, on average, a patient in a private hospital costs three times more than one in a public hospital.^13^ In particular, public hospitals have 1.4 ICU beds per 10 thousand inhabitants, while private hospitals have 4.9. This difference is more pronounced in the North and Northeast regions, with 0.9 and 1.5 beds per 10 thousand inhabitants in public hospitals against 4.7 and 5.5 beds per 10 thousand inhabitants for private hospitals, respectively.^14^ Complementary to a hospital’s funding is its level of strain. Our findings are in line with the findings of previous studies^15,16^ and suggest the importance of funding public hospitals and better managing the healthcare network, with profound implications for policy making in Brazil.

Finally, we showed how one can use the XCOVID-BR model in order to estimate the mortality risk of specific groups of the Brazilian population. In other words, one can apply XCOVID-BR to arbitrary sections of the Brazilian population and estimate the differential risk from COVID-19 (Figure 1), helping policy makers to take informed decisions regarding vaccination/non-pharmacological preventative measures, hospital management and healthcare network organization in an equitable way.

As an example, we showed how the risk distribution differs between two representative areas: The wealthier Paraná and more socioeconomically challenged Pernambuco (Figure 7). The variation in probability distributions is striking. Accounting for age, the dominant clinical predictor of mortality, it is apparent that the probability distribution is heavily skewed to higher probabilities of recovery in the wealthier (Paraná) region and this is particularly apparent in younger patients and in privately-funded hospitals.

The current vaccination plan proposed by the Brazilian Ministry of Health^17^ closely follows the plans devised by countries in Europe such as the UK.^18^ In particular, prioritization is mostly based on age and comorbidities. While these factors are undoubtedly significant, we have shown here that in Brazil they are not the sole risk factors and that socioeconomic and structural factors are actually as important in order to reduce COVID-19 mortality.^19–21^ We hope that our findings will help the Brazilian Ministry of Health adopt vaccination/non-pharmacological preventative measures that are properly tailored to the complex socioeconomic profile of Brazil.

Finally, given the changing nature of the virus, with ever more frequent emergence of SARS-CoV-2 variants, it is worth stressing the significance of data-driven risk factor discovery. Indeed, one expects that the relative importance of biological and structural COVID-19 risk factors depends on case fatality rate, transmissibility and response to vaccination efforts of the new variants. A data driven approach seems to be an agile approach to understand such an ever-changing scenario.

## Data Availability

The data and codes used for this work are made publicly available.

https://github.com/PedroBaqui/XCOVID-BR

## CONTRIBUTORS

PB and MvdS conceived the research question. All authors designed the study and analysis plan. PB performed the machine learning analysis. VM drafted the initial version of the manuscript. AE oversaw the clinical review of the methods and manuscript. All authors critically reviewed early and final versions of the manuscript.

## DATA SHARING

SIVEP-Gripe data are publicly available at opendatasus.saude.gov.br/dataset/bd-srag-2020. Our analysis code and XCOVID-BR are available at github.com/PedroBaqui/XCOVID-BR.

