## Supplementary materials for "Comparing COVID-19 risk factors in Brazil using machine learning: the importance of socioeconomic, demographic and structural factors"

#### CONTENTS

I. Geographical spread of SARS-CoV-2 1

II. Differences among the macro-regions 4

III. Machine learning details 5
A. Adopted features 5
B. Hyperparameters 5
C. Handling of missing values 5
D. Machine learning performance 5
E. Feature importance robustness 6

References 6

A. TRIPOD Statement 7

#### I. GEOGRAPHICAL SPREAD OF SARS-COV-2

Brazil is divided into 27 Federative Units (26 states and 1 federal district), which are grouped into five macro-regions:

- North: Rondônia (RO), Acre (AC), Amazonas (AM), Roraima (RR), Pará (PA), Amapá (AP), Tocantins (TO);
- Northeast: Bahia (BA), Piaui (PI), Maranhão (MA), Ceará (CE), Rio Grande do Norte (RN), Paraíba (PB), Pernambuco (PE), Alagoas (AL), Sergipe (SE);
- Central-West: Distrito Federal (DF), Goiás (GO), Mato Grosso (MT), Mato Grosso do Sul (MS);
- Southeast: São Paulo (SP), Rio de janeiro (RJ), Espírito Santo(ES), Minas Gerais (MG);
- South: Santa Catarina (SC), Paraná (PR), Rio Grande do Sul (RS).

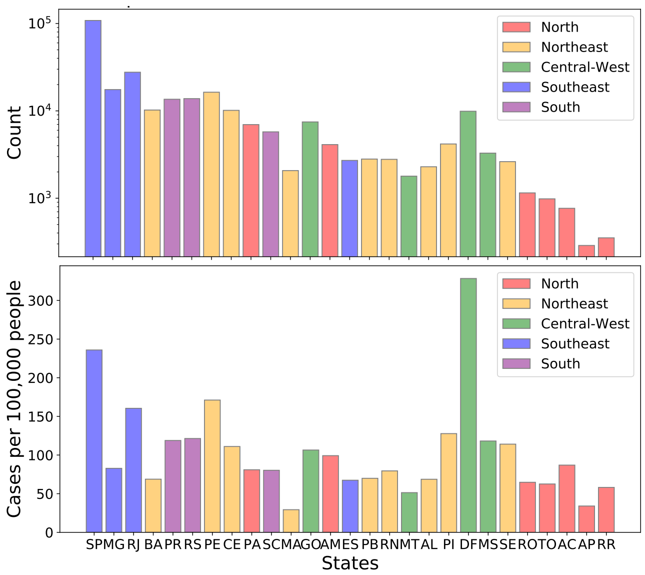


Figure 1. **Distribution of the 279,987 hospitalized patients with SARS-CoV-2 among Brazilian states according to absolute number of cases and number of cases per 100,000 people.** The different colors represent the Brazilian macro-regions. States are ordered according to their population, larger on the left.

Figure 1 shows the distribution of hospitalized SARS-CoV-2 patients, colored according to macro-region. By comparing the distribution of hospitalized SARS-CoV-2 patients among the Brazilian states with earlier investigations (Figure 1),[1] it is evident that the pandemic propagated though Brazil, affecting basically all the states. This indicates a failure in non-pharmaceutical interventions such as social distancing.

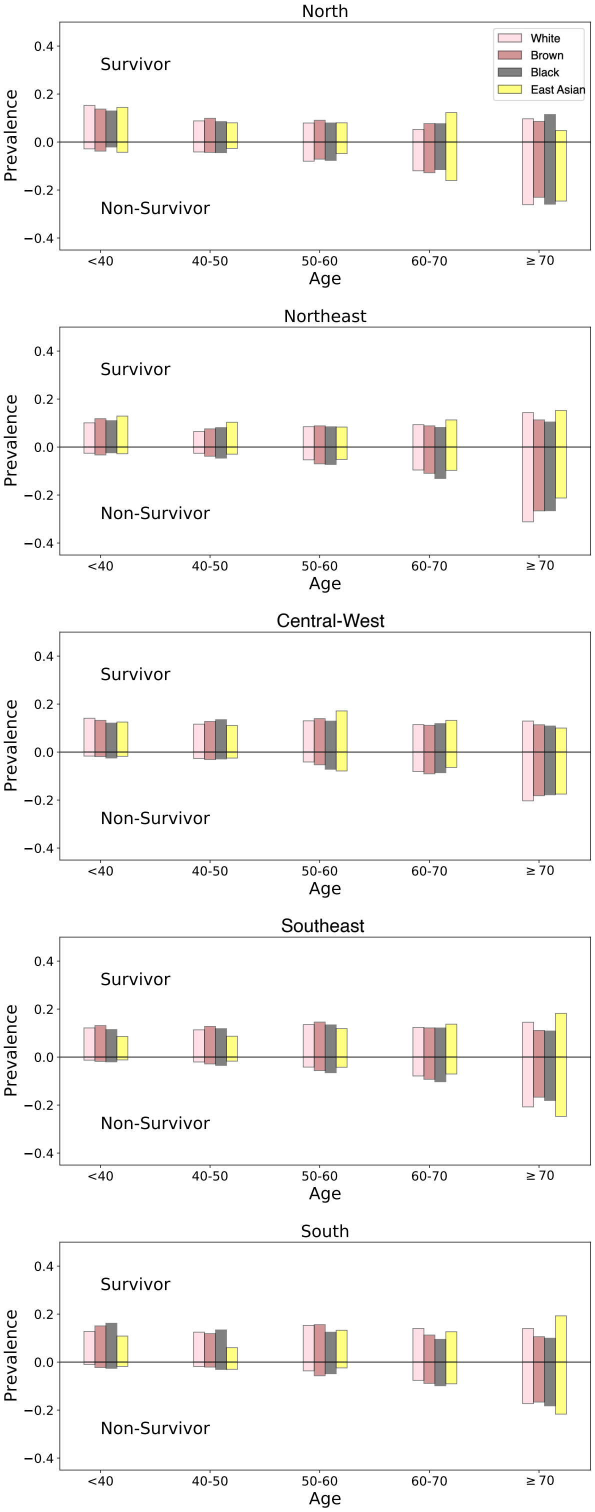


Figure 2. **Distributions of ethnicity according to age.** The normalization is such that all the fractions of a given ethnicity add to unity (to adjust for differences in ethnic prevalence). We exclude Indigenous patients for clarity because of their small numbers in the study population.

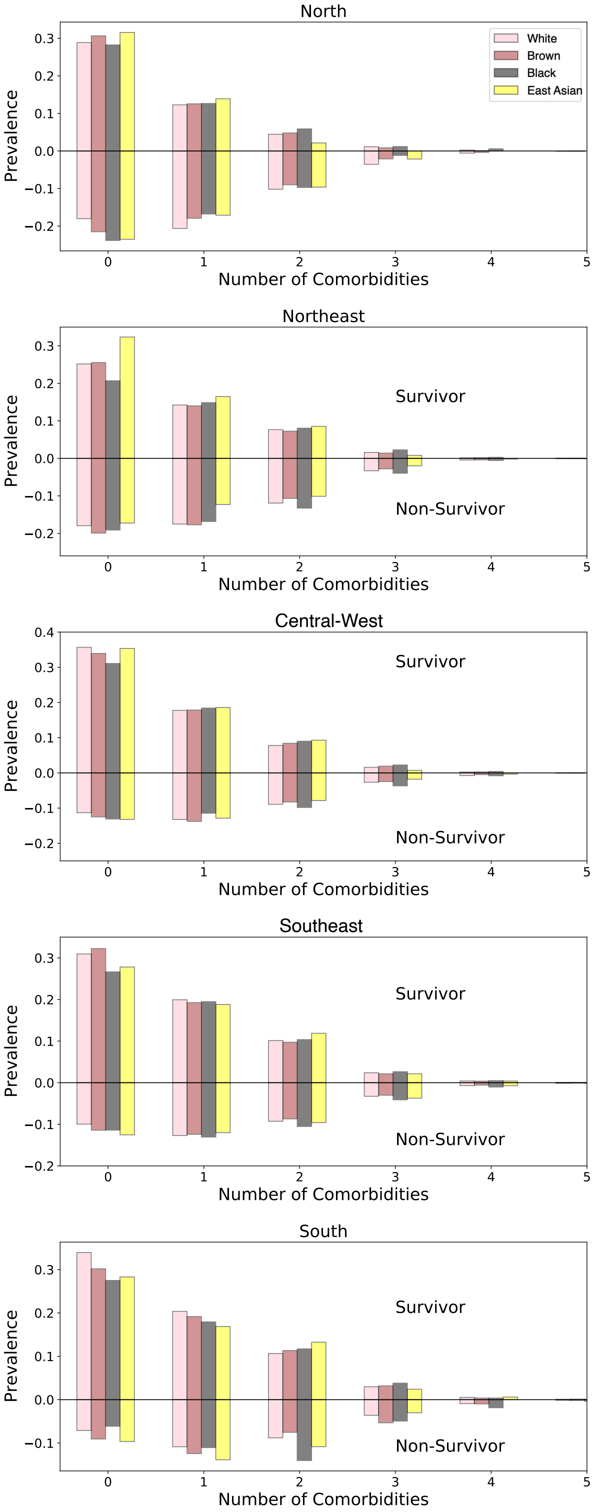


Figure 3. **Distributions of ethnicity according to number of comorbidities.** The normalization is such that all the fractions of a given ethnicity add to unity. We exclude Indigenous patients for clarity because of their small numbers in the study population.

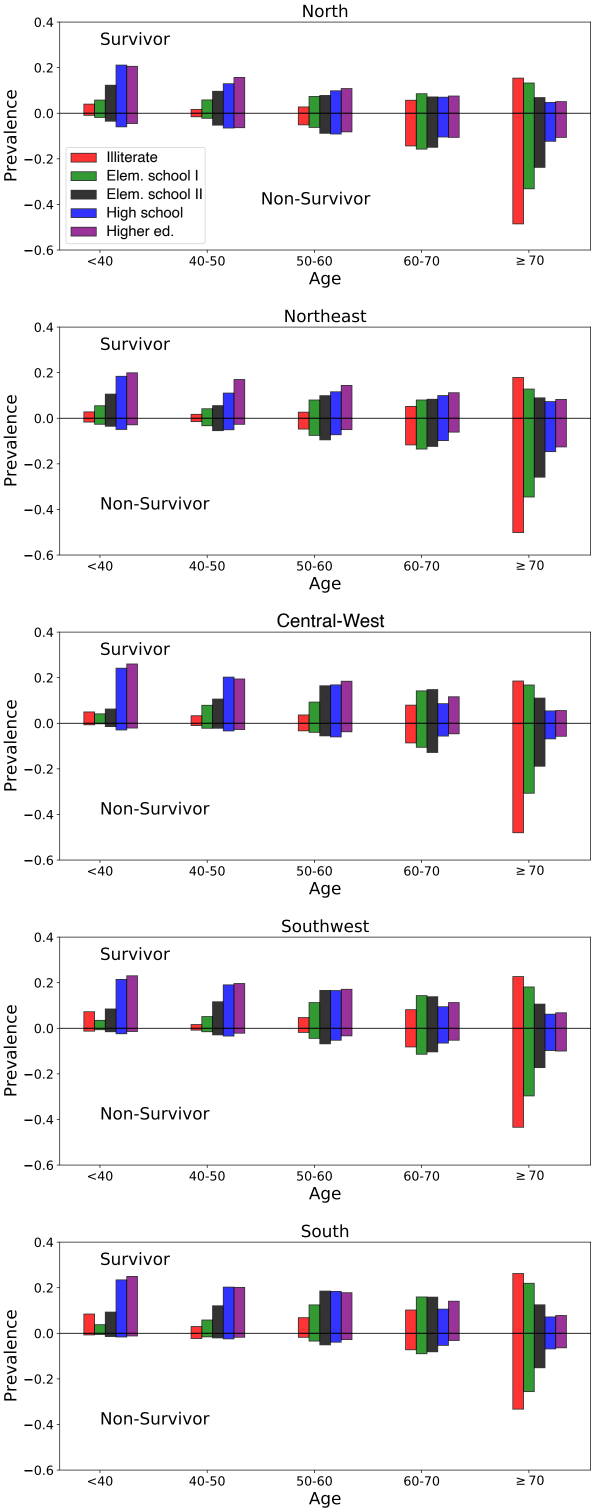


Figure 4. **Distributions of education level according to age.** The normalization is such that all the fractions of a given education level add to unity.

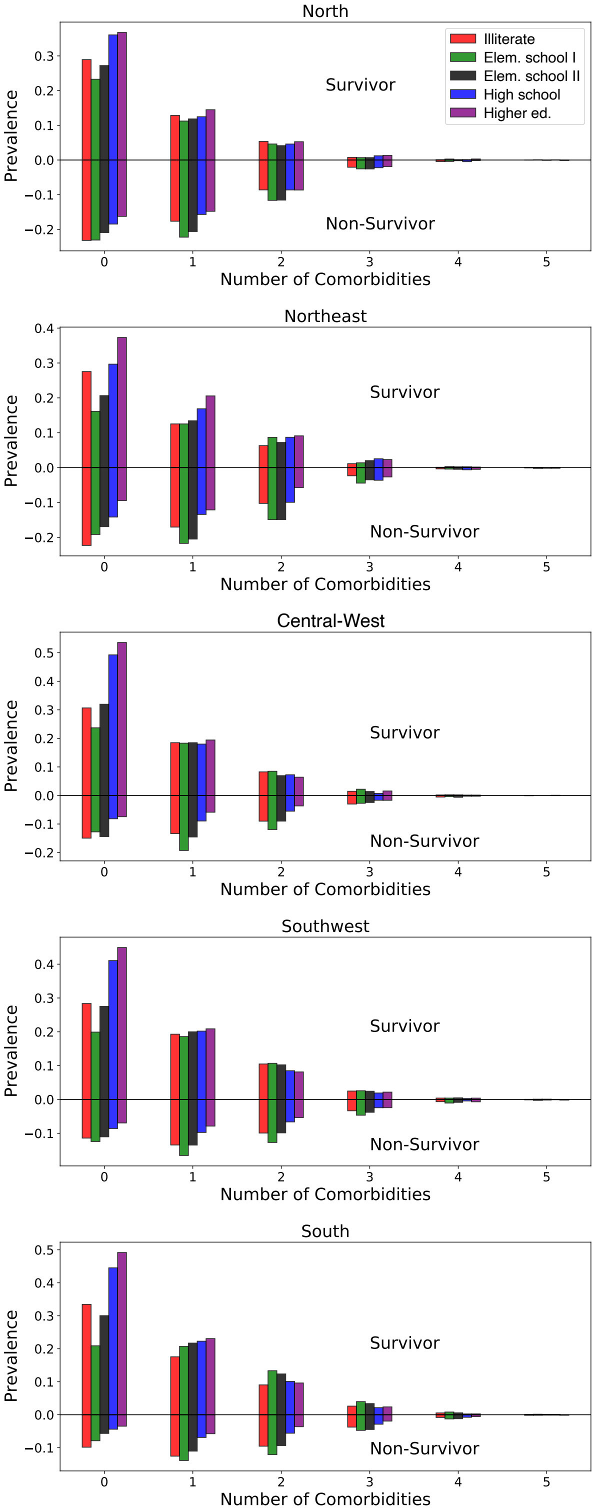


Figure 5. **Distributions of education level according to number of comorbidities.** The normalization is such that all the fractions of a given education level add to unity.

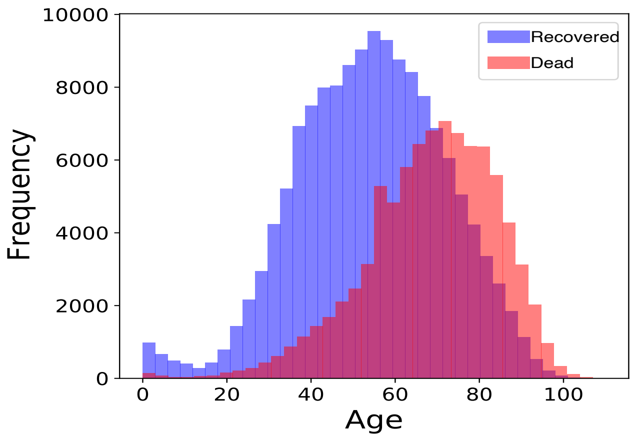


Figure 6. **Age distribution of the hospitalized patients considered in this work.**

#### II. DIFFERENCES AMONG THE MACRO-REGIONS

Brazil is a large and diverse country. Although each state has its individual particularities, each macro-region shares among its members socioeconomic similarities. Figures 2-5 give an overall view of the differences among the five Brazilian macro-regions. The common trend is that the South and Southeast regions have responded better to the pandemic compared to the North and Northeast regions. The figures are normalized so that the sum of each ethnicity is equal to one, independently. In this analysis we used data from 242,679 patients, relaxing some constraints on municipality and hospital data.

Figures 2-3 are stratified according to ethnicity as the latter is correlated with poverty.[2] Poverty implies a higher lever of susceptibility to SARS-CoV-2 as remote working may not be possible and poor people tend to live in crowed households with less access to sanitation.[3] Moreover, they do not have access to private health care. Figures 4-5 are stratified according to education, which is again correlated with poverty. Illiteracy refers to patients without education and more than seven years of age. As shown by Figure 6, our dataset includes mostly adult patients.

#### III. MACHINE LEARNING DETAILS

##### A. Adopted features

For completeness we list below the features that we considered when implementing the machine learning algorithms:

- **Clinical Factors**: age, sex, ethnicity, comorbidities (cardiovascular disease, liver disease, asthma, pulmonary disease, renal disease, hematologic disease, diabetes, obesity, neurological disease, immunosuppression, sum of comorbidities), symptoms (fever, vomiting, cough, sore throat, respiratory discomfort, shortness of breath, diarrhea, SpO2<95%, sum of symptoms).
- **Socio-Geographic**: education, state, MHDI, city type, distance to hospital.
- **Health System**: funding (private or public), strain.

In total we considered 30 features.

##### B. Hyperparameters

The hyper-parameters adopted for XGBoost are:

```
n_estimators: 200
eta: 0.2
max_depth: 4
gamma: 1
subsample: 0.9
colsample_bytree: 0.9
```

More at github.com/PedroBaqui/XCOVID-BR.

##### C. Handling of missing values

The SIVEP-Gripe catalog has missing values. In the case of comorbidities or symptoms we imputed missing values as the clinical feature being absent for the individual.[1] For the remaining variables we did not perform pre-processing for the XGB algorithm as the latter already imputes missing data. On the other hand, for the LR, KNN, NN, RF and SVM models we adopted scikit-learn's SimpleImputer. Table I shows the percentages of patients with missing values for all the features that we consider.

##### D. Machine learning performance

Table II shows the performance of the machine learning algorithms considered in this work. The XGBoost algorithm achieves an excellent score and is adopted in the analysis of the main text.

Table I. Percentages of patients with missing values.

| Feature | No. (%) |
|---|---|
| Age | 0 (0.0%) |
| Sex | 0 (0.0%) |
| Funding Model | 0 (0.0%) |
| MHDI | 0 (0.0%) |
| Ethnic group | 20877 (9.0%) |
| Macro-region | 0 (0.0%) |
| City type | 24353 (10.5%) |
| Education level | 65366 (28.3%) |
| Comorbidities | |
| Cardiovascular disease | 104841 (45.4%) |
| Asthma | 132906 (57.5%) |
| Diabetes | 112748 (48.8%) |
| Pulmonary disease | 131744 (57.0%) |
| Obesity | 131784 (57.0%) |
| Immunosuppression | 132993 (57.5%) |
| Renal disease | 131622 (57.0%) |
| Liver disease | 134321 (58.1%) |
| Neurological disease | 131494 (56.9%) |
| Hematologic disease | 133999 (58.0%) |
| Symptoms | |
| Fever | 21277 (9.2%) |
| Vomiting | 57379 (24.8%) |
| Cough | 18765 (8.1%) |
| Sore throat | 51474 (22.3%) |
| Respiratory discomfort | 34119 (14.8%) |
| Shortness breath | 20421 (8.8%) |
| Diarrhea | 54109 (23.4%) |
| SpO2 < 95% | 30981 (13.4%) |

| Model | AUC (95%CI) | $AP_{recovery}$ (95%CI) | $AP_{death}$ (95%CI) |
|---|---|---|---|
| **XGB** | **0.813** [0.810, 0.817] | **0.879** [0.876, 0.883] | **0.711** [0.704, 0.721] |
| **XGB***| **0.797** [0.793, 0.801] | **0.866** [0.863, 0.870] | **0.689** [0.682, 0.698] |
| RF | 0.798 [0.793, 0.803] | 0.867 [0.863, 0.871] | 0.686 [0.678, 0.696] |
| RF* | 0.781 [0.778, 0.786] | 0.854 [0.851, 0.859] | 0.661 [0.654, 0.669] |
| NN | 0.795 [0.791, 0.800] | 0.865 [0.861, 0.869] | 0.677 [0.670, 0.685] |
| NN* | 0.782 [0.779, 0.786] | 0.855 [0.852, 0.859] | 0.660 [0.653, 0.670] |
| LR | 0.766 [0.761, 0.770] | 0.840 [0.835, 0.845] | 0.632 [0.622, 0.640] |
| LR* | 0.763 [0.759, 0.768] | 0.837 [0.833, 0.842] | 0.629 [0.619, 0.639] |
| SVM | 0.766 [0.761, 0.770] | 0.842 [0.838, 0.847] | 0.635 [0.627, 0.644] |
| SVM* | 0.761 [0.757, 0.766] | 0.838 [0.834, 0.843] | 0.628 [0.619, 0.638] |
| KNN | 0.764 [0.760, 0.769] | 0.834 [0.830, 0.839] | 0.634 [0.627, 0.642] |
| KNN* | 0.751 [0.746, 0.756] | 0.825 [0.820, 0.829] | 0.615 [0.607, 0.622] |

Table II. **Performance of the machine learning algorithms considered in this work.** The analyses with "*" sign do not consider symptoms.

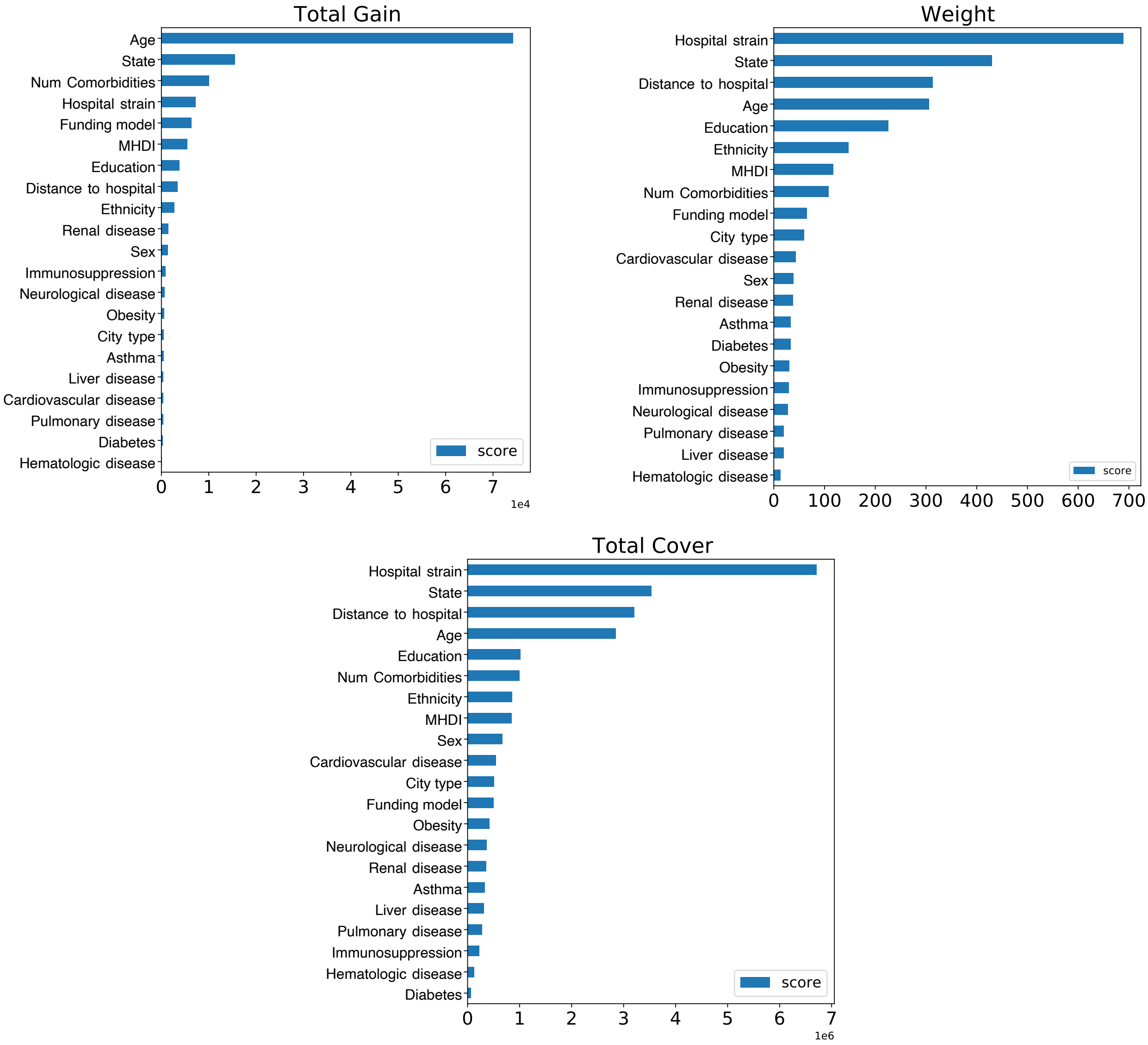


Figure 7. Different methods used to calculate feature importance for the XGB algorithm over the training set.

##### E. Feature importance robustness

The result of the feature importance analysis may depend on the specific method adopted. Here, in order to test the robustness of our results, we consider different feature importance methods for the XGB algorithm without symptom information.

The result is shown in Figure 7 which should be compared to Figure 4 of the main text. The *Total Gain* method adopts gain values associated with the features that divided the data. The *Weight* method uses the number of times a particular feature divided the data. Finally, the *Total Cover* method makes use of the number of data points affected by the cut where a feature was used. The *Permutation* method adopted in Figure 4 of the main text randomly breaks the relationship between feature and target and measures the decrease in the metric used.[4]

We note that the various methods all give a higher importance to socio-geographical and hospital-specific features compared to comorbidities. In particular, hospital strain is confirmed to be a very important factor.

[1] Baqui P, Bica I, Marra V, Ercole A, and van der Schaar M. Ethnic and regional variations in hospital mortality from COVID-19 in Brazil: a cross-sectional observational study. *The Lancet Global Health*, 2020; **8**: 1018–26.

[2] IBGE. Desigualdades Sociais por Cor ou Raça no Brasil, 2019. biblioteca.ibge.gov.br/visualizacao/livros/liv101681_informativo.pdf. (accessed July 27, 2020).

[3] Tavares F and Betti G. Vulnerability, Poverty and COVID-19: Risk Factors and Deprivations in Brazil, 2020. researchgate.net/publication/340660228_Vulnerability_Poverty_and_COVID-19_Risk_Factors_and_Deprivations_in_Brazil (accessed May 10, 2020).

[4] xgboost developers. Python API Reference, 2020. xgboost.readthedocs.io/en/latest/python/python_api.html. (accessed August 23, 2020).

[5] Collins GS, Reitsma JB, Altman DG, and Moons KG. Transparent Reporting of a multivariable prediction model for Individual Prognosis Or Diagnosis (TRIPOD): The TRIPOD Statement. *Annals of Internal Medicine*, 2015; **162**: 55–63.

#### Appendix A: TRIPOD Statement

The study was conducted and reported in line with the Transparent Reporting of a multivariable prediction model for Individual Prognosis Or Diagnosis (TRIPOD).[5] The TRIPOD Statement is attached to the Supplementary Materials.

### TRIPOD Checklist: Prediction Model Development

| Section/Topic | Item | Checklist Item | Page |
|---|---|---|---|
| **Title and abstract** | | | |
| Title | 1 | Identify the study as developing and/or validating a multivariable prediction model, the target population, and the outcome to be predicted. | 1 |
| Abstract | 2 | Provide a summary of objectives, study design, setting, participants, sample size, predictors, outcome, statistical analysis, results, and conclusions. | 1 |
| **Introduction** | | | |
| Background and objectives | 3a | Explain the medical context (including whether diagnostic or prognostic) and rationale for developing or validating the multivariable prediction model, including references to existing models. | 1-2 |
| | 3b | Specify the objectives, including whether the study describes the development or validation of the model or both. | 1-2 |
| **Methods** | | | |
| Source of data | 4a | Describe the study design or source of data (e.g., randomized trial, cohort, or registry data), separately for the development and validation data sets, if applicable. | 2-3 |
| | 4b | Specify the key study dates, including start of accrual; end of accrual; and, if applicable, end of follow-up. | 2-3 |
| Participants | 5a | Specify key elements of the study setting (e.g., primary care, secondary care, general population) including number and location of centres. | 2-3 |
| | 5b | Describe eligibility criteria for participants. | 2-3 |
| | 5c | Give details of treatments received, if relevant. | -- |
| Outcome | 6a | Clearly define the outcome that is predicted by the prediction model, including how and when assessed. | 2-3 |
| | 6b | Report any actions to blind assessment of the outcome to be predicted. | -- |
| Predictors | 7a | Clearly define all predictors used in developing or validating the multivariable prediction model, including how and when they were measured. | 2-3 |
| | 7b | Report any actions to blind assessment of predictors for the outcome and other predictors. | -- |
| Sample size | 8 | Explain how the study size was arrived at. | 2-3 |
| Missing data | 9 | Describe how missing data were handled (e.g., complete-case analysis, single imputation, multiple imputation) with details of any imputation method. | 2-3 |
| Statistical analysis methods | 10a | Describe how predictors were handled in the analyses. | 2-3 |
| | 10b | Specify type of model, all model-building procedures (including any predictor selection), and method for internal validation. | 2-3 |
| | 10d | Specify all measures used to assess model performance and, if relevant, to compare multiple models. | 2-3 |
| Risk groups | 11 | Provide details on how risk groups were created, if done. | 2-3 |
| **Results** | | | |
| Participants | 13a | Describe the flow of participants through the study, including the number of participants with and without the outcome and, if applicable, a summary of the follow-up time. A diagram may be helpful. | 3 |
| | 13b | Describe the characteristics of the participants (basic demographics, clinical features, available predictors), including the number of participants with missing data for predictors and outcome. | 3-4 |
| Model development | 14a | Specify the number of participants and outcome events in each analysis. | 3-4 |
| | 14b | If done, report the unadjusted association between each candidate predictor and outcome. | -- |
| Model specification | 15a | Present the full prediction model to allow predictions for individuals (i.e., all regression coefficients, and model intercept or baseline survival at a given time point). | 3 |
| | 15b | Explain how to the use the prediction model. | 3 |
| Model performance | 16 | Report performance measures (with CIs) for the prediction model. | 4 |
| **Discussion** | | | |
| Limitations | 18 | Discuss any limitations of the study (such as nonrepresentative sample, few events per predictor, missing data). | 5-6 |
| Interpretation | 19b | Give an overall interpretation of the results, considering objectives, limitations, and results from similar studies, and other relevant evidence. | 5-6 |
| Implications | 20 | Discuss the potential clinical use of the model and implications for future research. | 5-6 |
| **Other information** | | | |
| Supplementary information | 21 | Provide information about the availability of supplementary resources, such as study protocol, Web calculator, and data sets. | 6 |
| Funding | 22 | Give the source of funding and the role of the funders for the present study. | 6 |

We recommend using the TRIPOD Checklist in conjunction with the TRIPOD Explanation and Elaboration document.
